# Cyclic exit strategies to suppress COVID-19 and allow economic activity

**DOI:** 10.1101/2020.04.04.20053579

**Authors:** Omer Karin, Yinon M. Bar-On, Tomer Milo, Itay Katzir, Avi Mayo, Yael Korem, Boaz Dudovich, Eran Yashiv, Amos J. Zehavi, Nadav Davidovitch, Ron Milo, Uri Alon

## Abstract

Many countries have applied lockdown that helped suppress COVID-19, but with devastating economic consequences. Here we propose exit strategies from lockdown that provide sustainable, albeit reduced, economic activity. We use mathematical models to show that a cyclic schedule of 4-day work and 10-day lockdown, or similar variants, can prevent resurgence of the epidemic while providing part-time employment. The cycle pushes the reproduction number R below one by reduced exposure time and by exploiting the virus latent period: those infected during work days reach peak infectiousness during lockdown days. The number of work days can be adapted in response to observations. Throughout, full epidemiological measures need to continue including hygiene, physical distancing, compartmentalization, testing and contact tracing. This conceptual framework, when combined with other interventions to control the epidemic, can offer the beginnings of predictability to many economic sectors.

---

Current non-pharmaceutical interventions to suppress COVID-19 use testing, contact tracing, physical distancing, mask use, identification of regional outbreaks, compartmentalization down to the neighborhood and company level, and population-level quarantine at home known as lockdown^1–4^. The aim is to flatten the infection curve and prevent overload of the medical system until a vaccine becomes available.

Lockdown is currently in place in many countries. It has a large economic and social cost, including unemployment on a massive scale. Once a lockdown has reduced the number of critical cases to a desired goal, a decision must be reached on how to exit it responsibly. The main concern is the risk of resurgence of the epidemic. One strategy proposes reinstating lockdown when a threshold number of critical cases is exceeded in a resurgence, and stopping lockdown again once cases drop below a low threshold^2,5,6^ (Fig 1A,S1). While such an “adaptive triggering” strategy can prevent healthcare services from becoming overloaded, it leads to economic uncertainty and continues to accumulate cases with each resurgence.

**Fig. 1.**
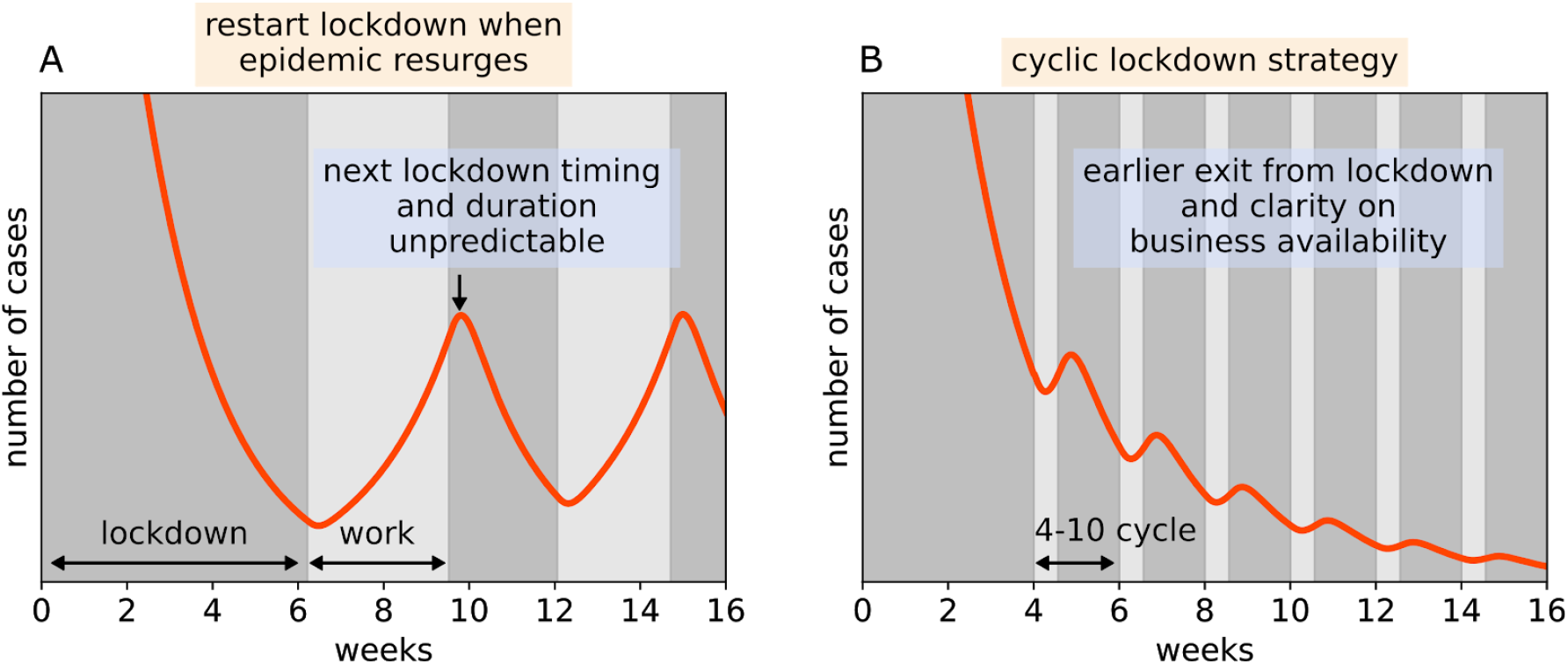
Cyclic work-lockdown strategy can suppress the epidemic, prevent resurgence and offer predictable part-time employment. *a) Exit from lockdown carries the risk of resurgence of the epidemic, with need to re-enter prolonged lockdown. b) A cyclic work-lockdown strategy prevents resurges by keeping the average R<1. It thus allows an earlier exit from lockdown, and provides a clear part-time work schedule. Transmission rates provide R in lockdown and work days of R*_*L*_ = 0.6 *and R*_*W*_ = 1.5 *respectively*.

Here we carefully propose an exit strategy from lockdown that can prevent resurgence of the epidemic while allowing sustained, albeit reduced, economic activity. The strategy can be implemented when lockdown has succeeded in stabilizing the number of daily critical cases to a value that the health system can support. Hereafter when we say ‘lockdown’ we mean population-level quarantine at home, together with all other available interventions such as testing and social distancing.

The basic idea is to keep the effective reproduction number R, defined as the average number of people infected by each infected individual, below 1. When R is below 1, the number of infected people declines exponentially, a basic principle of epidemiology.

To reduce R below 1, we propose a cyclic schedule with k continuous days of work followed by n continuous days of lockdown. As shown below, 4 days of work and 10 days of lockdown is a reasonable cycle that allows a repeating 2-week schedule. Epidemiological measures should be used and improved throughout, including rapid testing, contact isolation and compartmentalization of workplaces and regions. The cyclic strategy can thus be considered as a component of the evolving policy toolkit that can be combined with other interventions.

By “work days” we mean release from lockdown with strict hygiene and physical distancing measures on the same k weekdays for everyone. It can include the entire population including schools, except for quarantined infected individuals and people in risk groups who may be in quarantine. More conservatively, it can include workers in selected sectors of the economy. Remote work should be encouraged for sectors that can work from home.

A staggered cyclic strategy is also predicted to be effective, in which the population is divided into two sets of households that work on alternating weeks^7^, each with a k-work:(14-k)-lockdown schedule (Fig 2). The staggered strategy has the advantage that production lines can work throughout the month and transmission during workdays is reduced due to lower density, whereas the non-staggered strategy has the advantage that lockdown days are easier to enforce.

**Fig. 2.**
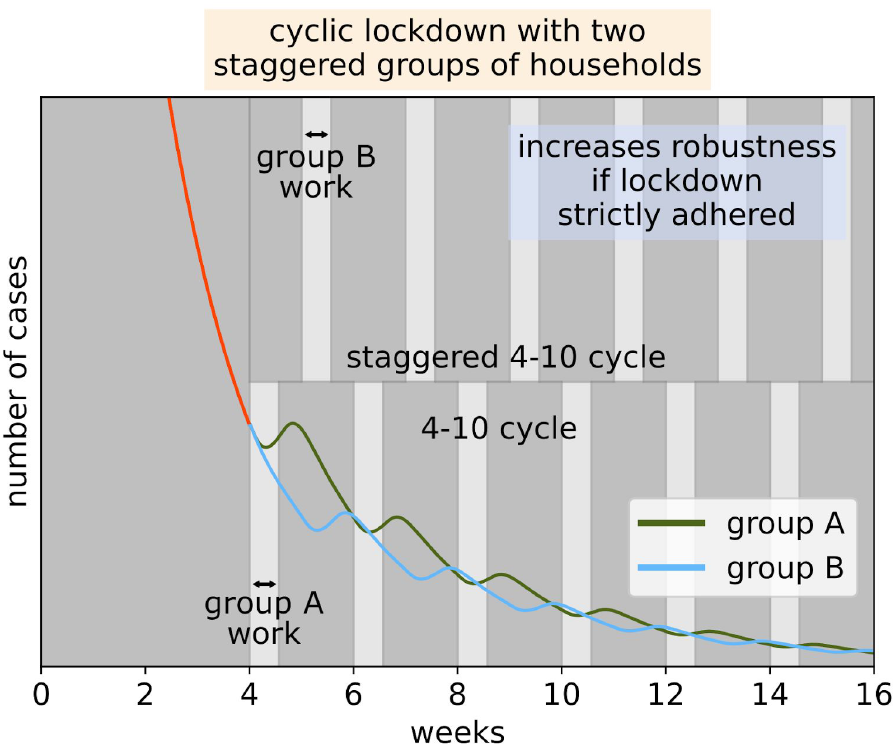
Staggered cyclic work-lockdown strategy in which the population is divided into two groups of households that work on alternating weeks. *Shown is I(t) from the SEIR-Erlang deterministic model with mean latent period of 3 days and mean infectious period 4 days* ^*8*^. *Transmission rates in lockdown and work give R*_*L*_ = 0.6 *and R*_*W*_ = 1.5 *respectively. Density compensation is* ϕ = 1.5 *and non-compliance is 10% (see Methods)*.

The cyclic strategies reduce the mean R by two effects: *restriction* and *anti-phasing*. The restriction effect is a reduction in the time T that an infectious person is in contact with many others, compared to the situation with no lockdown. For example, a 4-day work: 10-day lockdown cycle reduces T to 2/7 T ≈ 0.3T.

The anti-phasing effect uses the timescales of the virus against itself (Fig. 3). Most infected people are close to peak infectiousness for about 3-5 days, beginning ≈3 days after being exposed^9,10^. A proper work-lockdown cycle, such as a 4-work:10-lockdown schedule, allows most of those infected during work days to reach maximal infectiousness during lockdown, and thus avoid infecting many others. Those with symptoms can be infectious for longer ^10^, but can be detected by their symptoms and remain hospitalized, isolated or (self-)quarantined along with their household members, preventing secondary infections outside the household.

**Fig. 3.**
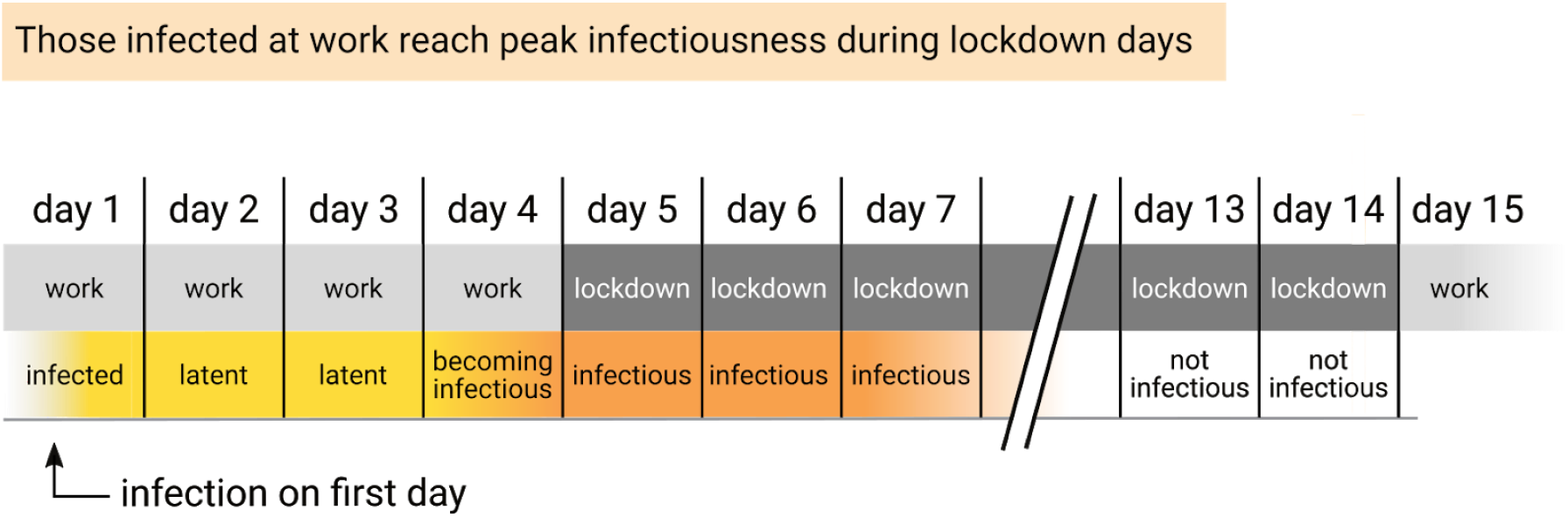
The cyclic exit strategy is aided by placing peak infectiousness in the lockdown days. SARS-CoV-2 has an average latent (non-infectious) period of about 3 days. A 14-day cycle in which people enter lockdown after 3 or 4 work days benefits from this property. Even those infected on the first day of work spend most of their latent period at work and reach peak infectiousness during lockdown. This reduces the number of secondary infections.

The staggered cyclic strategy can further reduce mean R by reducing density during work days, leading to lower transmission rates (Methods).

The cyclic strategy can be synergistically combined with rapid testing and contact isolation. Household-level testing at the end of the lockdown period and before return to work or school can help shorten infection chains.

Simulations using a variety of epidemiological models, including SEIR models and stochastic network-based simulations, show that a cyclic strategy can suppress the epidemic provided that the lockdown is effective enough (Fig. 4, Table 1). A 4-10 cycle seems to work well for a range of parameters and is robust to uncertainties in the model (Fig. S2, S3). In these simulations, the transmission parameters during work days and lockdown days can be described by the effective replication numbers that describe extended periods of work and lockdown conditions, *R*_*W*_ and *R*_*L*_, respectively. If lockdown is as strong as strong as in some chinese cities, with estimated *R* _*L*_ ≈ 0.3 _11,12_, a 4-10 cycle suppresses the epidemic even if workday *R*_*W*_ is as large as in the early days of the epidemic in Europe, with *R*_*W*_ ≈ 3 − 4 _13_. A weaker lockdown, with *R* _*L*_ ≈ 0.6 − 0.8, as currently estimated in several European countries (April 20, 2020)^14^ can support a cyclic strategy with 2-4 work days when strict measures are enforced during workdays providing *R*_*W*_ ≈ 1.5 − 2 (Table 1). Simulation on social networks provides a similar range of conditions of a cyclic strategy to control the epidemic (Fig S5). Ideally, measures will eventually bring R during workdays down below 1, as in South Korea’s control of the epidemic in early 2020, making lockdown unnecessary.

**Table 1.** **Effective replication numbers for a 4-10 cyclic strategy in several scenarios. R_W_ and R_L_ are the replication numbers that would be observed in continuous periods of work and lockdown, respectively. Parameters are of Fig 1**,**3**.

| $R_W$ | $R_L$ | Effective replication number , $Re$, in 4-10 cycle ($Re < 1$ means epidemic declines) | Effective replication number , $Re$, in 4-10 cycle with two staggered groups |
| --- | --- | --- | --- |
| 1.5 | 0.6 | 0.86 | 0.85 |
| 2.0 | 0.6 | 0.94 | 0.93 |
| 2.5 | 0.6 | 1.03 | 1.02 |
| 2.5 | 0.3 | 0.8 | 0.8 |

**Fig. 4.**
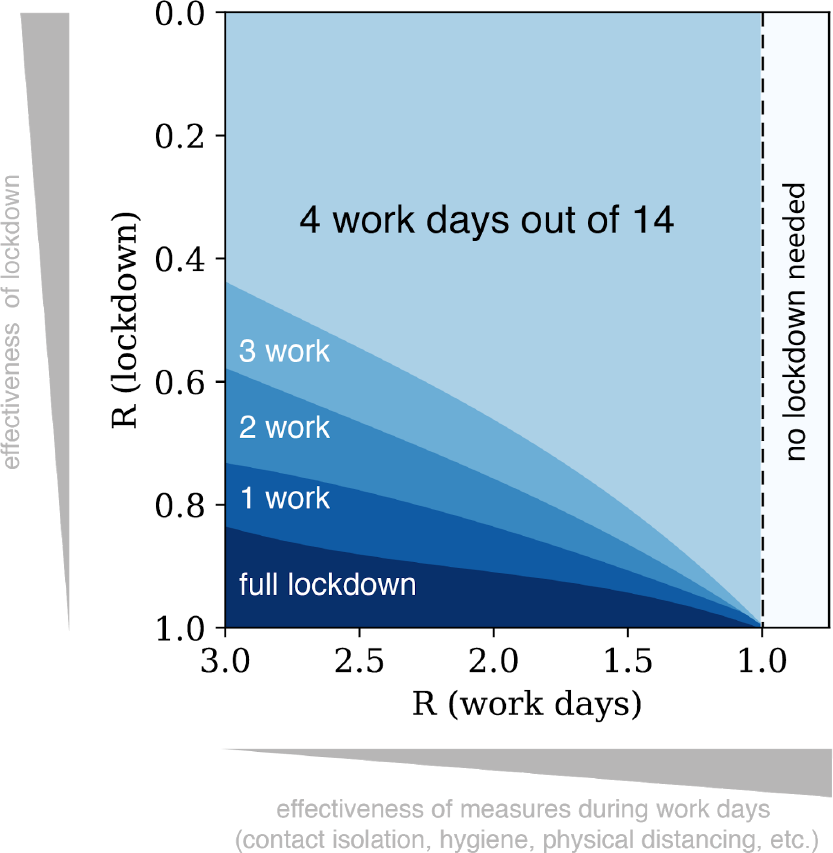
Cyclic strategy with k workdays and 14-k lockdown days controls the epidemic for a range of effective replication numbers at work and lockdown. Each region shows the maximal number of work days in a 14-day cycle that provide decline of the epidemic. Simulation used a SEIR-Erlang deterministic model with mean latent period of 3 days and infectious periods of 4 days. Results are robust to uncertainty in model parameters (Fig S2).

An important consideration is that the cyclic strategy is adaptive, and can be tuned when conditions change and the effects of the approach are monitored. For example, weather conditions may affect R^5,6^, as well as advances in regional monitoring and contact tracing. If one detects, for example, that a 4:10 strategy leads to an increasing trend in cases, one can shift to a cycle with fewer work days. Conversely, if a strong decreasing trend is observed, one can shift to more work days and gain economic benefit (Fig 5). A conservative approach can exit lockdown with 1 day of work, build up to two days, and so on.

**Fig. 5.**
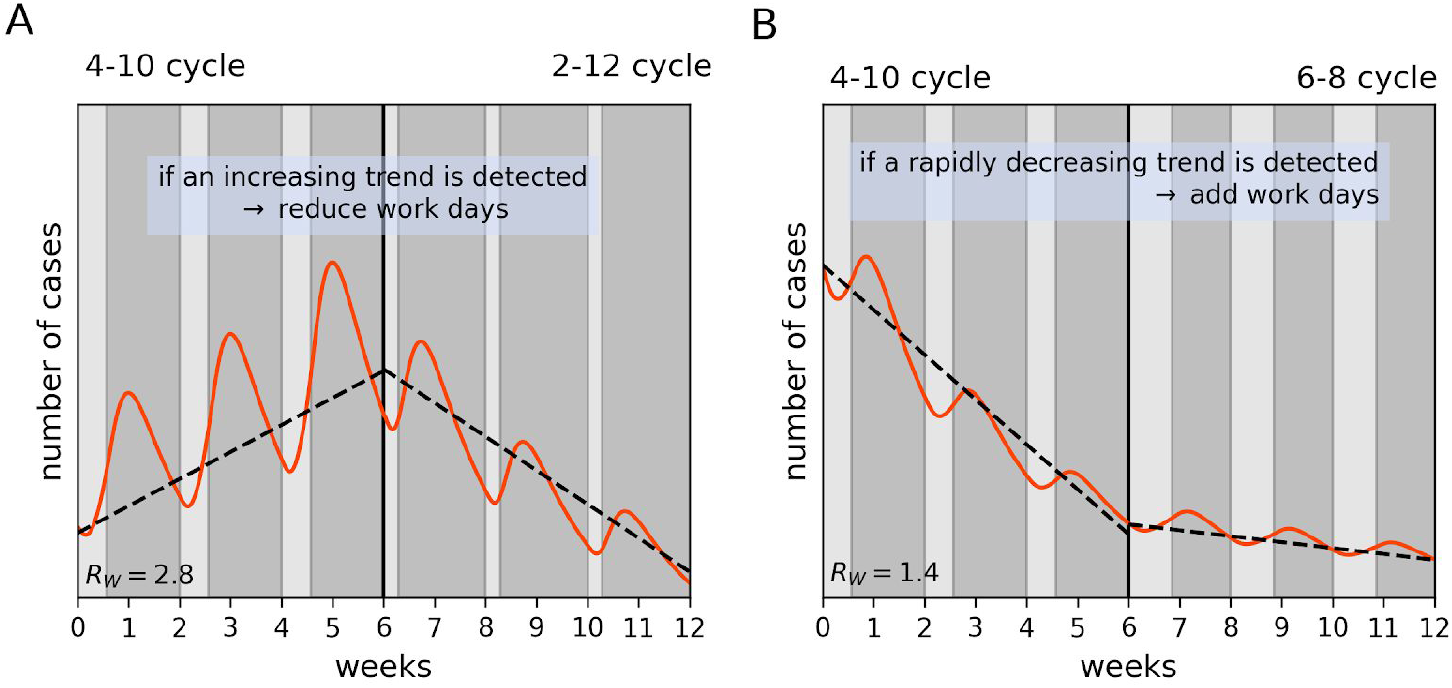
The cyclic strategy can be tuned according to the trends in case numbers over weeks. (a) If average R is above 1, cases will show a rising trend, and number of work days in the cycle can be reduced to achieve control. (b) Number of work days per cycle can be increased when control meets a desired health goal.

**Fig. 6.**
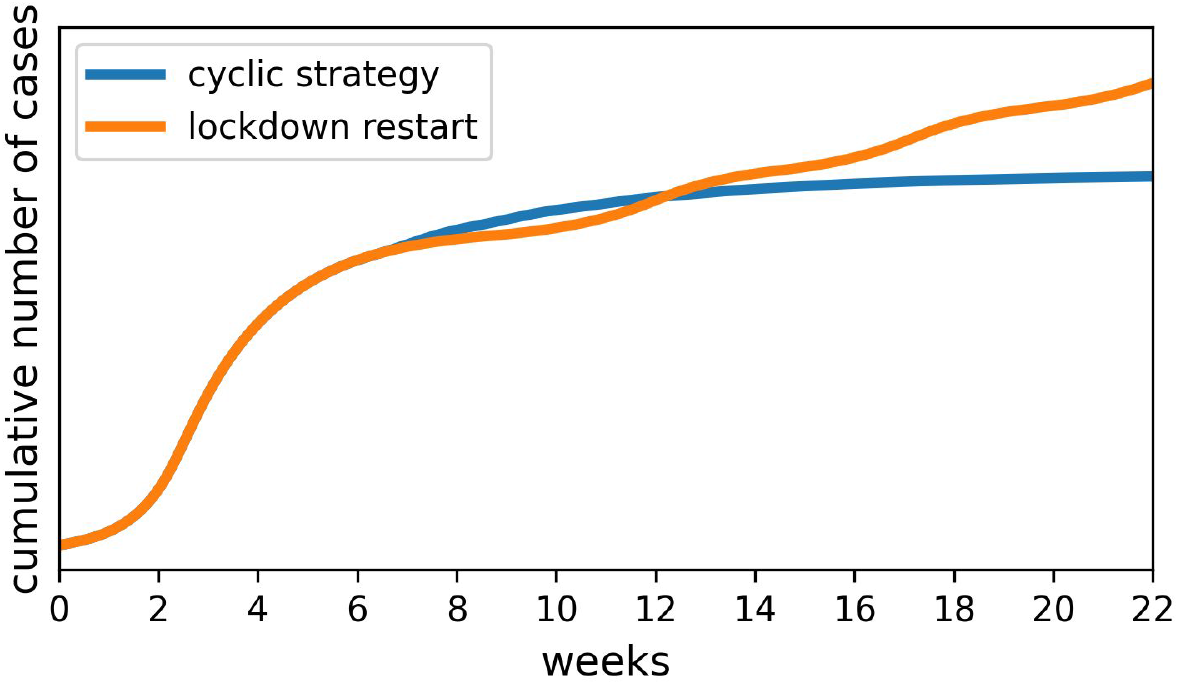
The cumulative number of cases under a cyclic strategy is lower at long times than in a strategy that restarts lockdown when the epidemic resurges. Cumulative number of cases is shown for the simulations of Fig 1a (red) and Fig1b (blue). Even though exit from lockdown is earlier in the cyclic strategy case, the cumulative number of cases associated with this strategy is lower in the long term than the accumulated cases when lockdown is released and then restarted once the epidemic resurges. The relative benefit of the cyclic strategy is further increased by considering non-COVID-19-related health consequences of extended lockdown during resurgences, alleviated by the cyclic strategy.

Measures will be required during the work days to ensure that people do not excessively compensate for the lockdown periods by having so many more social connections that R is strongly increased. This may include sound epidemiological measures such as the continuation of banning large social events and clear communication campaigns by the health authorities to enhance adherence to hygiene and physical distancing. Shared work spaces can be disinfected to reduce future infection through surfaces. Extensive rapid testing and contact tracing should be developed and extended in parallel^15^.

The economic benefits of a cyclic strategy include part-time employment to millions who have been put on leave without pay or who have lost their jobs. This mitigates massive unemployment and business bankruptcy during lockdown. Prolonged unemployment during lockdown and the recession that is expected to follow can reduce worker skill and carries major societal drawbacks. Unemployment also has detrimental health effects which include exacerbation of existing physical and mental illnesses. High levels of unemployment have been associated with increases in morbidity and mortality ^16^.

The cyclic strategy offers a measure of economic predictability, potentially enhancing consumer and investor confidence in the economy which is essential for growth and recovery. It can also be equitable and transparent in terms of who gets to exit lockdown.

For these reasons, a cyclic strategy can be maintained for far longer than continuous lockdown. This allows time for developing a vaccine, treatment, rapid testing and buildup of herd immunity without overwhelming health care capacity.

The cyclic strategy does not seem to have a long-term cost in terms of COVID-19 cases compared to a start-stop lockdown policy triggered by resurgences. Comparing the two strategies shows that in the mid-term and long term, the start-stop strategy accumulates more cases due to resurgences (Fig. 5). This does not depend heavily on parameters: the fundamental reason is that new cases arise during each resurgence. Thus, a strategy that restarts lockdown with every resurgence uses feedback to effectively keep average R close to 1, and continues to accumulate cases. In contrast, the cyclic strategy keeps average R below one, and thus prevents resurgences.

The cyclic strategy can apply at many scales: to a company, a school, a town or an entire country. Regions or organizations that adopt this strategy are predicted to resist infections from the outside. An infection entering from the outside cannot spread widely because average R<1. After enough time, if this is applied globally, there is even a possibility for the epidemic to be eradicated, in the absence of mutations or unknown reservoirs.

The cyclic strategy can work in regions with insufficient testing capacity, as long as the lockdown phases provide low enough transmission. This may apply to a large part of the earth’s population.

The exact nature of the intervention can be tuned to optimize economy and minimize infection. It can be tested for a limited duration such as a month, and in a limited region. The cyclic strategy can be synergistically combined with other approaches to suppress the epidemic and address the economic crisis.

## Methods

### SEIR model

The deterministic SEIR model is *dS* /*dt* =− β*SI, dE* /*dt* = β*SI* − σ*E, dI* / *dt* = σ*E* − γ*I*, where S,E and I are the susceptible, exposed (noninfectious) and infectious fractions. Parameters calibrated for COVID-19 ^8^ are [ineq, and S=1 is used to model situations far from herd immunity. The values used for β are defined in each plot where *R* = β/ γ. The analytical solution for cyclic strategies is in (SI).

### SEIR-Erlang model

The SEIR model describes an exponential distribution of the lifetimes of the exposed and infectious compartments. In reality these distributions show a mode near the mean. To describe this, we split E and I into two artificial serial compartments each with half the mean lifetime of the original compartment ^17^. This describes Erlang-distributed lifetimes (the distribution of the sum of two exponentially distributed random variables) with the same mean transition rates as the original SEIR model. Thus, *dS*/*dt* =− β*SI, dE*_1_/*dt* = β*SI* − 2σ*E*_1_, *dE*_2_ /*dt* = 2σ*E*_1_ − 2σ*E*_2_, *dI*_1_ /*dt* =2σ*E*_2_ − 2γ*I*_1_, *dI*_2_ /*dt* = 2γ*I*_1_ − 2γ*I*_2_, where *I* = *I*_1_ + *I*_2_, and *R* = β/ γ. In the figures we used a worst-case assumption of no herd immunity, namely *S* ≈ 1. Herd immunity further reduces case numbers. Case numbers are in arbitrary units, and can describe large or small outbreaks. The deterministic simulation describes a fully-mixed population. Population structure typically reduces overall outbreak peak size ^18^ compared to a fully mixed situation with the same mean transmission rate, but includes the possibility of high attack rates in certain sub-populations.

### Staggered cyclic strategy, SEIR-Erlang model

We model two groups, A and B, with a susceptible, exposed, infectious and removed compartment for each group. The SEIR-Erlang model for group A is:

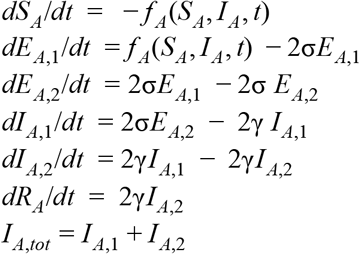

and an analogous equation for group B. We assume that each group consists of half of the population. This causes density at work to be reduced ^7^. For ease of comparison to the non-staggered case, we refer to the replication numbers of a single fully mixed population with a cyclic strategy, namely *R* = *R*_*W*_ on work days and *R* = *R*_*L*_ during lockdown. In the staggered case, during lockdown, as opposed to work, individuals from a group interact primarily with their own household. The density in the household is not affected by dividing the population into two staggered work groups. Hence, the effective R remains *R*_*L*_ .

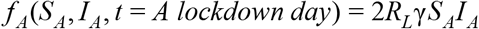

Where the factor of 2 normalizes *S*_*A*_ = 0.5. During work days, we can estimate the number of transmissions at work and not at home by *R*_*W*_ − *R*_*L*_. This gives the following equation:

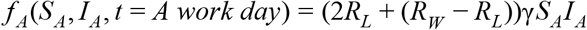

With analogous equations for group B.

We next model cross-transmission between the groups. Due to the expected difficulty of enforcing a staggered work schedule as compared to a non-staggered cycle strategy, we assume a leakage term due to a fraction of individuals ρ from each group that does not adhere to their lockdown. These non-adherers instead interact with the other group during the other groups’ work days.

When group B is in lockdown, infectious non-adherers from group B can infect individuals from group A who are in their work days. This rate is modeled as proportional to the replication number for people infected at work and not at home, *R*_*W*_ − *R*_*L*_ :

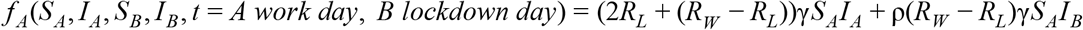

When individuals from group A are in lockdown and non-adhere, they can be infected from individuals from group B on group B work days. We also add a higher-order term for susceptible ‘leaking’ individuals from group A that meet non-adherent infectious ‘leaking’ individuals from group A during group A lockdown:

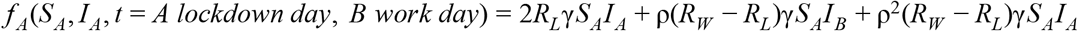

When both groups are in lockdown at the same time, there is no leakage:

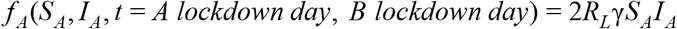

Note that for complete leakage ρ = 1 and under symmetry assumptions *I*_1_ = *I*_2_, the equations become identical to the case of a single fully mixed population:

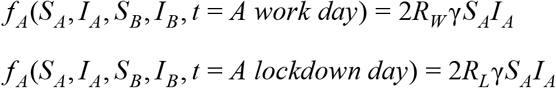

So far, we assumed that density at work is half that of the non-staggered case. However, in practice, compensatory mechanisms might lead to a higher effective density. For example, people might cluster to maintain a level of social interaction, or certain work-day situations may require a fixed density of individuals. These effects can be modelled by adding a density compensation parameter ϕ which rescales the work-day infection rate. This number is ϕ = 2 for complete compensation of infectivity where density at work is not affected by partitioning, or ϕ = 1 is the staggered model above with half the density at work. We obtain the following equations:

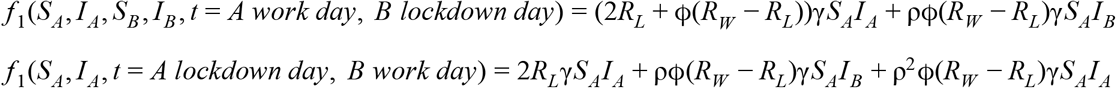

With analogous equations for group B.

### Stochastic SEIR model on social networks with epidemiological measures

We also simulated a stochastic SEIR process on social contact networks. Each node i represents an individual and can be in a susceptible, exposed, infected or removed state (i.e. quarantined, recovered or dead). Lifetime in the E and I states is drawn from an Erlang distribution of shape 2 with means *T*_*E*_ and *T* _*I*_. The total infectivity of a node β_*i*_ is drawn from a long tailed distribution to account for super-spreaders. The probability of infection per social link j, *q*_*ij*_, is set either constant for all links connected to node i or drawn from an exponential distribution to account for heterogeneity in infection rates. Node states are updated at each time step. Network models include Erdos-Renyi and small world networks. During lockdown, a fraction of the links are inactivated (same links for each lockdown phase).

### Linearity of transmission risk with exposure time

In order for restriction of exposure time to be effective, probability of infection must drop appreciably when exposure time is reduced. This requires a low average infection probability per unit time per social contact, q, so that probability of infection,*p* = 1 − *exp*(− *qT*), does not come close to 1 for exposure time T on the order of days. For COVID-19, an infected person infects on the order of R=3 people on average during the infectious period of mean duration D=4 days. If the mean number of social contacts is C, which is estimated at greater than 10, one has q∼DR/C<0.1/day. Thus infection probability on the scale of hours to a few days is approximately linear with exposure time: 1 − *exp*(− *qT*) ≈ *qT*. This is consistent with the observation that infected people do not typically infect their entire household, and with the linearity observed in influenza transmission^19^. We also tested a scenario using network models in which some contacts have much higher q than others (exponentially distributed q between links). A mildly lower R in lockdown is required to provide a given benefit of the cyclic strategy than when q is the same for all links.

**Fig. S1.**
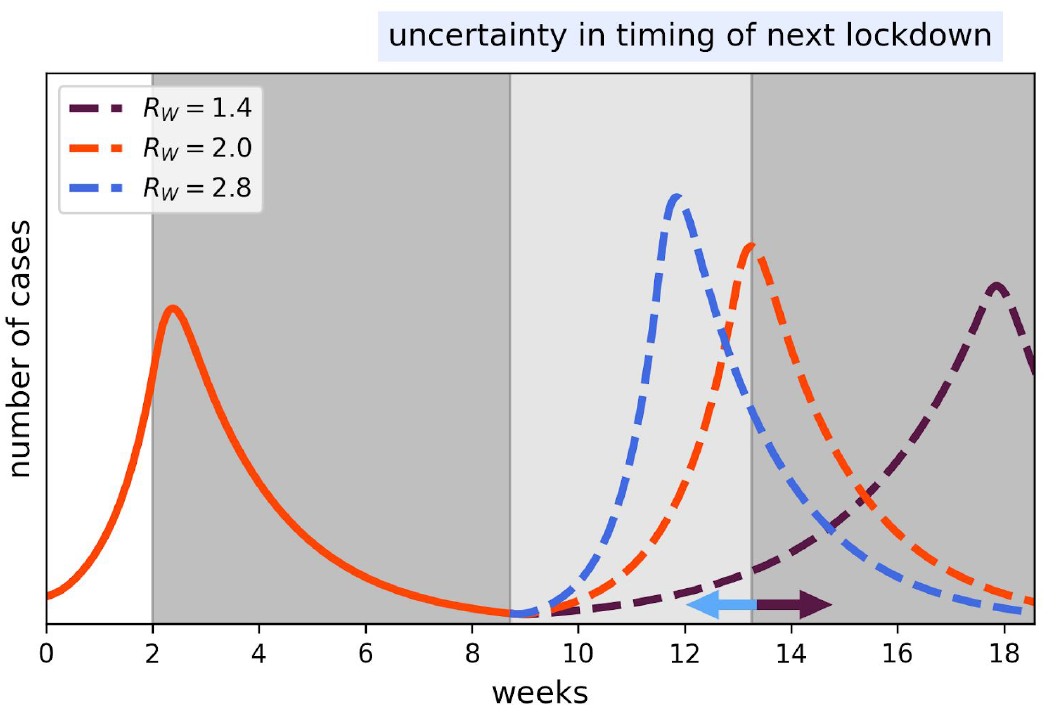
Reinstating lockdown with each resurgence leads to uncertainty in the timing of new lockdown. SEIR-Erlang model simulation showing the initial growth phase of an epidemic in the first two weeks, triggering a lockdown of 7 weeks. Lockdown is reinstated once a threshold of cases is exceeded. We show three scenarios with different effective reproduction numbers after lockdown is first lifted (R_W_=1.4, R_W_=2.0 and R_W_=2.8), leading to a wide distribution of the time at which the case threshold is crossed and lockdown is reinstated.

**Fig S2.**
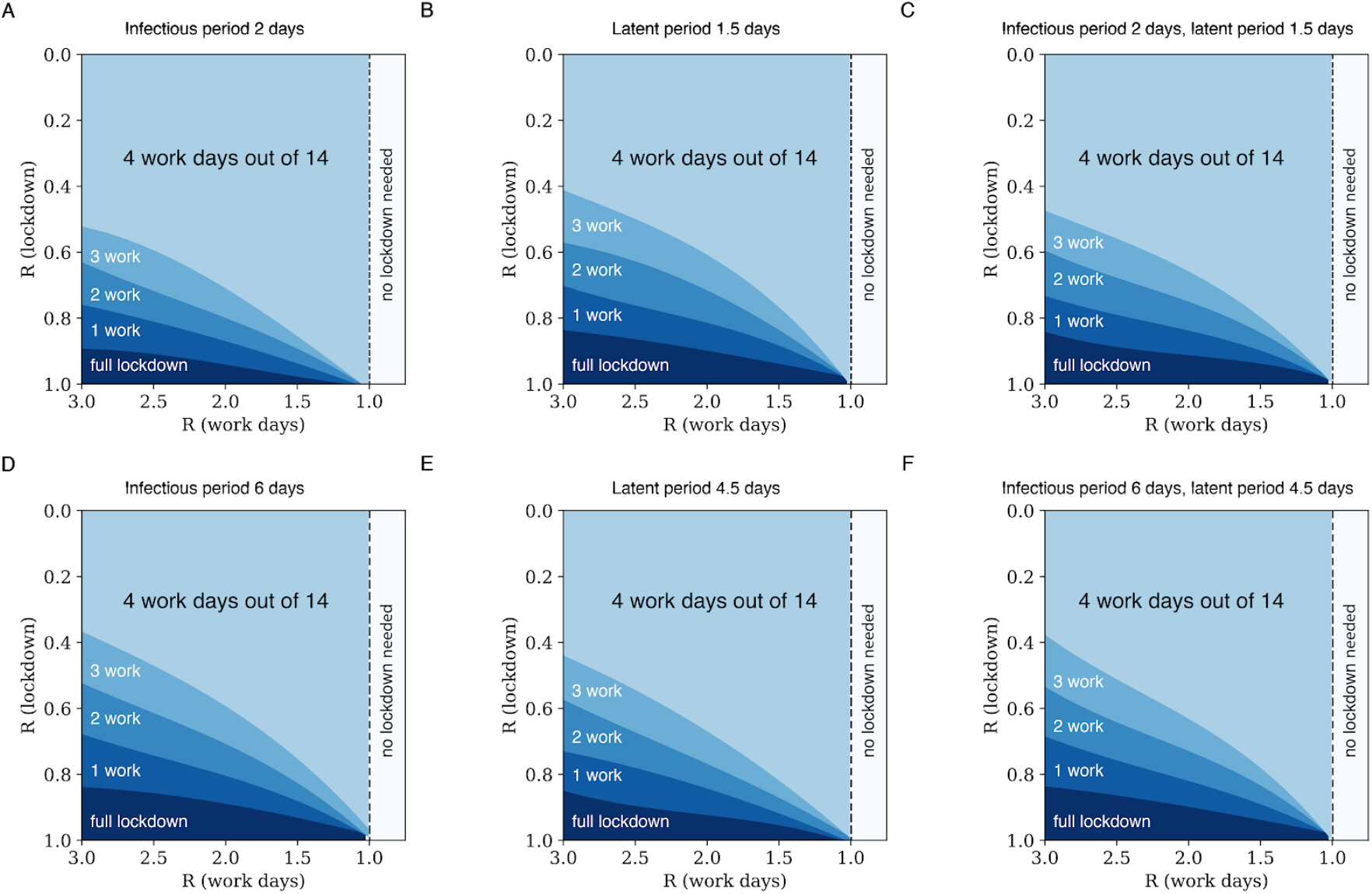
The cyclic strategy is insensitive to variations in the model parameters. *The SEIR-Erlang model has two free parameters, the lifetimes of the latent and infectious periods, given by T*_*E*_ = 1/σ *and T* _*I*_ = 1/γ. *The reference parameters used in the main text are T*_*E*_*=3 days, and T*_*I*_*=4 days based on the COVID-19 literature*^8^. *The panels show the regions in which effective R<1 with (A) T*_*E*_*=3d and T*_*I*_*=2d, (B) T*_*E*_*=1*.*5d and T*_*I*_*=4d, (C) T*_*E*_*=1*.*5d and T*_*I*_*=2d, (D) T*_*E*_*=3d and T*_*I*_*=6d, (E) T*_*E*_*=4*.*5 d and T*_*I*_*=4d, and (F) T*_*I*_*=6d, T*_*E*_*=4*.*5 d. These and similar parameter variations make small differences to the phase plots*.

**Fig. S3.**
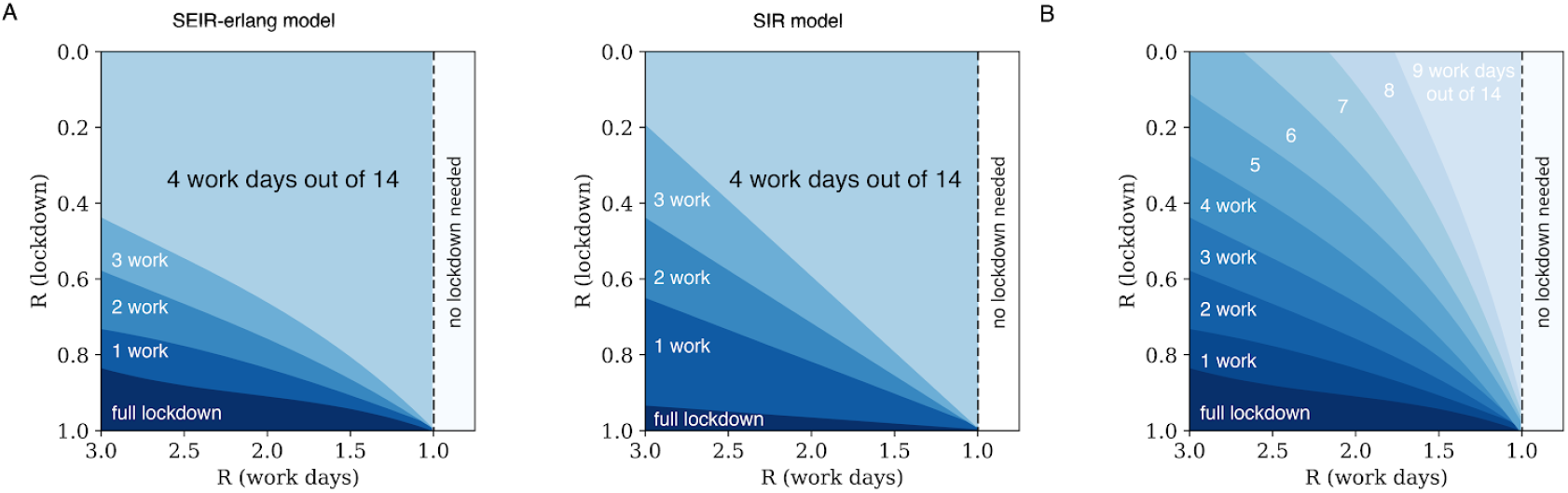
A SIR deterministic model captures some of the effects. *(A) The SIR model (right panel) lacks the exposed (non-infectious) compartment. It shows that the cyclic lockdown strategies can control the epidemic, but at smaller parameter regions for each given strategy than the SEIR-Erlang model (left panel). The difference is biggest at large ratios of R at work and lockdown, where SEIR-Erlang has an advantage. The SIR model is dS*/*dt* =− β*SI, dI*/*dt* = β*SI* − γ*I. The replication number is R* = β/ γ. *Here* γ = 1/7 *day*^−1^ *(the plot is unaffected by this parameter), and S=1. Effective R in the SIR model is the average R weighted by the fraction of time for work and lockdown. For analytical work on optimal epidemic control in the SIR model see [https://osf.io/rq5ct/]. Note that the axes in this plot are inverted with respect to Fig 3. (B) In this figure, each region shows the maximal number of workdays where the epidemic can be suppressed for the SEIR erlang model, for work days in the range of 0-11*.

**Fig. S4.**
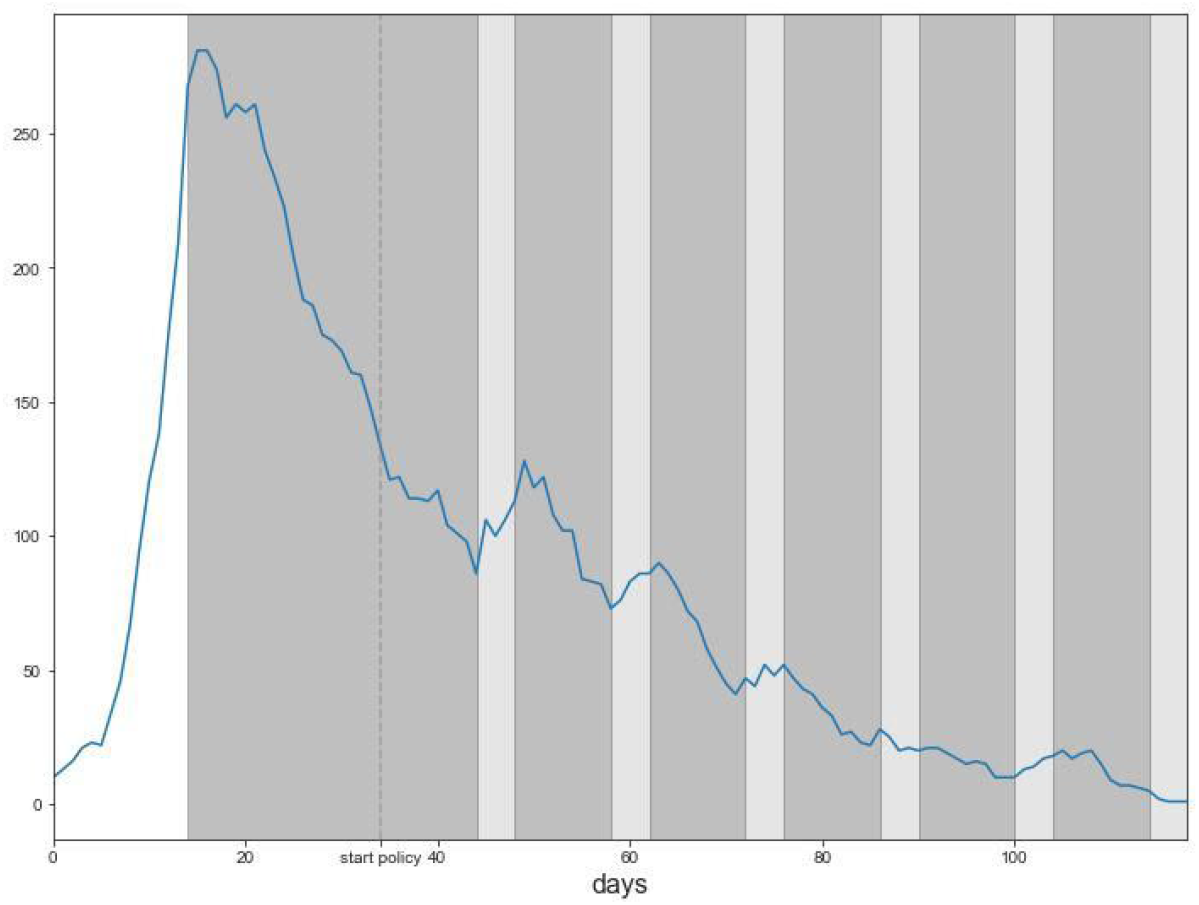
Stochastic simulation of a 4-work-10-lockdown cyclic strategy using a SEIR process simulated on a contact network. Infected nodes versus time from a simulation run using the SEIRSplus package from the Bergstrom lab, https://github.com/ryansmcgee/seirsplus. Contact network has a power-law-like degree distribution with two exponential tails, with mean degree of 15, N=10^4 nodes, sigma=gamma=1/3.5 days, beta=0.95 till day 14, lockdown beta=0.5 with mean degree 2 (same edges removed every lockdown period), work day beta=0.7. Probability of meeting a non-adjacent node randomly at each timestep instead of a neighbor node is p=1 before day 14, in lockdown p=0, workday p=0.3. Testing is modeled to quarantine 1% of infected nodes per day, with no contact tracing. Shaded regions are lockdown periods, light gray regions are workdays. Initial conditions were 10 exposed and 10 infected nodes.

**Fig S5.**
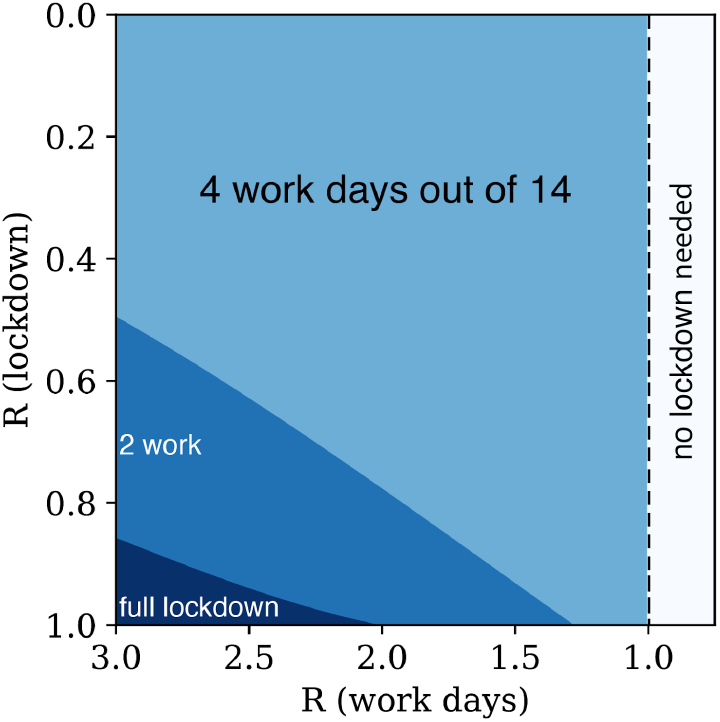
A stochastic simulation on a small-world network shows a similar range in which epidemic is controlled by a cyclic strategy. *A custom simulator uses a stochastic SEIR process on a social network. The social network is small-world with with N* = 10^4^, *mean degree C=16 and fraction of long-range connections probability p* = 1 − *R*_*L*_ /*R*_*W*_. *Timesteps are one day. In work days transmission occurs along edges, with q*_*W*_ = *R*_*W*_ /(*C T* _*inf*_ *). In lockdown days, the long range links of each node are inactivated (the same links are inactivated every day), with remaining links signifying the household. Transitions between exposed, infectious and removed states are determined by Erlang (shape=2) distributed times determined for each node at the beginning of the simulation. The effective R*_*W*_, *R*_*L*_ *were inferred for each condition by simulating continuous work and continuous full lockdown conditions*.

**Fig S6.**
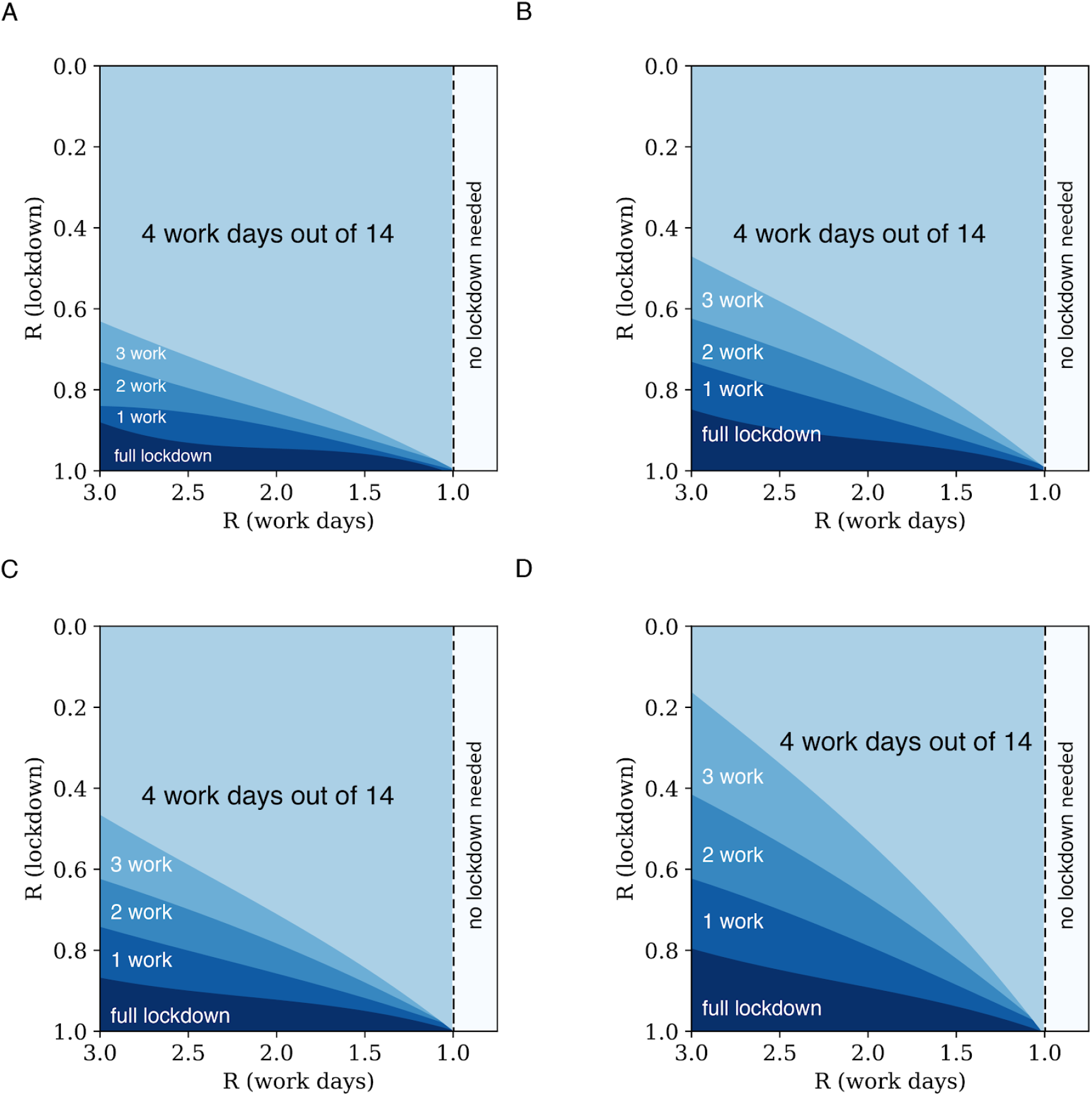
Staggered cyclic strategies control the epidemic for various degrees of density compensation and non-compliance. Each region shows the maximal number of work days in a 14-day cycle that provide decline of the epidemic. Simulation used a SEIR-Erlang deterministic model with mean latent period of 3 days and infectious periods of 4 days. Density compensation φ and non-compliance (cross transmission) η parameters were as follows: (a) φ=1,η=0.1 (b) φ=1.5,η=0.1 (c) φ=1,η=0.3 (d) φ=1.5,η=0.3. Code can be found at https://github.com/omerka-weizmann/2_day_workweek.

### SI section: Economic perspective on cyclic lockdown strategies

We summarize work on the economic repercussions of the COVID-19 pandemic and its attendant lockdown policies. Then, and in light of this discussion, we discuss some of the economic implications of the adaptive cyclic exit strategy.

Pandemics and the policy reaction to them constitute a real, substantial, and sudden supply shock to the economy. The following economic mechanisms can be outlined, based on empirical studies of past pandemics and of COVID 19:

a. They engender a substantial fall in GDP and its uses. Analyses of the economic consequences of the 1918 Spanish flu (Great Influenza Pandemic) found that the influenza led to 39 million deaths (which translate to 150 million deaths in 2020 terms), a drop in GDP of 6%, and a consumption decline of 8% in typical countries. These losses are comparable to those seen in the last global Great Recession of 2008-2009^20^. In fact, the expected annual cost of pandemic influenza falls in the same range as does that of climate change between 0.2% and 2% of global income. A pandemic that exceeds 720,000 deaths per year worldwide would cost $570 billion per year ($80 billion in lost income and $490 billion in mortality cost), 0.7% of global income^21^.
b. Longer term costs may emerge via hysteresis effects, whereby a temporary rise in unemployment, engendered by the fall in GDP, generates a permanently higher one^22,23^. This may entail loss of skill by the unemployed over time, leading them to be “unemployable”^24^.
c. Following the decline in supply, demand declines too. Thus consumption and investment decline^25^ is a recent treatment of the mechanism applied to the current context). It is to be expected that fiscal demand policy will respond in terms of health policy expenditures, disaster relief, provision of funds to liquidity-constrained households and firms, and general support of aggregate demand^22,25^.
d. The pandemic may lead to disruption in financial markets, including the banking system and stock markets.

In the current paper, which deals with an exit strategy from lockdown policies, the economic mechanism underlying the effects of such policies is particularly important. It was recently analyzed, for COVID 19, by Kaplan, Moll, and Violante^26^. A key idea in this analysis is that the replication number is a function of economic variables, including consumption of “social” goods and market work (labor). The social good is one that involves many social interactions, such as going to a restaurant. Economic outcomes under lockdown policies are determined by the operation of at least two margins: (i) how flexible is each job in terms of working remotely; and (ii) whether production is intense in the social good (i.e., has high inputs of the social good) or in the regular consumption good. The authors give the example of waiters and shop assistants jobs as inflexible and high on social goods; in contrast, the production of a software engineer or an architect is highly flexible and not intense in the social good. Lockdown policy and economic behavior affect the transmission rate β{t} and hence the epidemic dynamics and its economic costs, with dependence on the cited two margins. The mechanism is as follows: the initial effects of the epidemic and lockdown policy affect production and consumption; depending on how consumption and production of the social good are affected, β{t} is affected in turn; hence, epidemic dynamics are affected, and in turn influence production and consumption once more, and the process is iterated. The authors aim at modelling the emerging tradeoff between health and economic outcomes, given policy choices under this mechanism. They conjecture that there may be a region where both health and economic outcomes could be improved. The current proposal of a cyclic strategy aims at such improvement.

Lockdown policies have exerted a heavy toll in COVID19. A nowcast estimate for France by Institut National de la Statistique et des Etudes Economiques (INSEE) placed output decline in

March 2020, when France was on tight lockdown, at 35 percent below normal.^[i]^ An estimate by the UK Office for Budget Responsibility put the second quarter UK GDP decline at 35%^[ii]^. Forecasts by the IMF,^[iii]^ central banks and investment banks speak of declines in the whole of 2020, including a recovery phase, of 6%-12% of GDP. This is based on fairly optimistic scenarios of lockdown phase-out and resumption of normal activity.

The cyclic exit strategy from these lockdowns, proposed here, has at least two major types of effects:

First, it provides for a partial release from the lockdown, enabling more production and consumption. This has positive effects in terms of predictability, production projections and sharing, consumption planning, work in the informal sector, and more. For countries or regions that have a high share of the informal sector, the cyclic strategy can make lockdown days more bearable and hence enhance adherence.

It does have some disadvantages. For example, not all economic sectors may benefit similarly from the scheme. Thus, social good sectors that heavily rely on human contact such as flights, hotels, or restaurants are less likely to operate smoothly under this scheme, unless they make pronounced adjustments. However, the staggered version of the cyclic scheme alleviates some such potential problems, relating to disruptions in production continuity. The staggered strategy has the advantage that production can work throughout the month and transmission during workdays is reduced due to lower density, whereas the non-staggered strategy has the advantage that lockdown days are easier to enforce.

Second, by the dephasing effect that takes advantage of the latent period of the virus, it engenders relatively good health outcomes. At the same time it has beneficial non-Covid related health effects. For example, lack of medical testing such as colonoscopies or mammography, and disruptions in treatment, such as for blood pressure, end up costing lost life years. Lack of routine medical maintenance, such as dentistry and family medicine, end up creating even more damage. Likewise, for lack of mental health treatment, suicides, and more. Cyclic strategy can relieve these costs by lessening the disruptions. It should be recalled that such big-scale economic events, involving deep recessions, may lead to deaths of despair; see the recent comprehensive review by Case and Deaton^27^, who have done ground-breaking work on this issue.

The cyclic strategy has the potential to lighten the harsh tradeoff faced by policymakers. The essential mechanisms here are as follows: policy and economic behavior influence R{t} and worker participation in employment. Health and economic outcomes thus depend on the interaction between three dynamic paths:

a. R{t}
b. Worker employment.
c. Stop-start policy on lockdowns, dependent on I and on ICU capacity.

Better economic outcomes will be achieved the lower is the path of R{t} (point a), the higher is employment (point b), and the fewer lockdown resumptions there are (point c).

In this setup the cyclic k,14-k strategy has the following expected effects:

a. It increases economic activity, as it releases part of the lockdown.
b. While it does not fully improve COVID-19 outcomes (as it has periods of work with higher R) in the short term, it improves them in the mid-long term by preventing resurgences (Fig 5). This improves the tradeoffs faced by policymakers.
c. Any comparison to actual lockdowns depends on the relations between the three dynamic paths above in the cyclic strategy and in the actual policy strategy. In certain cases of these relations, the cyclic strategy can even improve health outcomes relative to existing lockdown outcomes.
d. The latent period idea serves to optimally determine k by minimizing the effective β{t} or R{t}. Hence it has effects operating to lower the path of R{t}, point a above.

To conclude, the cyclic strategy, utilizing the latent period of the coronavirus to optimize the cycle of work and lockdown, is a way to significantly ameliorate the health-economic tradeoffs faced by policymakers.

## Data Availability

All data supplied in the submitted file

## Footnotes

[i] See https://www.insee.fr/en/statistiques/4473305?sommaire=4473307#titre-bloc-13

[ii] See https://obr.uk/coronavirus-reference-scenario/#economic-scenario

[iii] See https://www.imf.org/en/Publications/WEO/Issues/2020/04/14/weo-april-2020

## Notes

### Competing Interest Statement

The authors have declared no competing interest.

### Funding Statement

None

## References

1. Flaxman, S. et al. Report 13: Estimating the number of infections and the impact of non-pharmaceutical interventions on COVID-19 in 11 European countries. (2020) doi: 10.25561/77731.

2. Ferguson, N. et al. Report 9: Impact of non-pharmaceutical interventions (NPIs) to reduce COVID19 mortality and healthcare demand. (2020).

3. Wang, C. J., Ng, C. Y. & Brook, R. H. Response to COVID-19 in Taiwan: Big Data Analytics, New Technology, and Proactive Testing. JAMA (2020) doi: 10.1001/jama.2020.3151.

4. Chen, S., Yang, J., Yang, W., Wang, C. & Bärnighausen, T. COVID-19 control in China during mass population movements at New Year. Lancet 395, 764–766 (2020).

5. Kissler, S. M., Tedijanto, C., Lipsitch, M. & Grad, Y. Social distancing strategies for curbing the COVID-19 epidemic. (2020) doi: 10.1101/2020.03.22.20041079.

6. Kissler, S. M., Tedijanto, C., Goldstein, E., Grad, Y. H. & Lipsitch, M. Projecting the transmission dynamics of SARS-CoV-2 through the postpandemic period. Science (2020) doi: 10.1126/science.abb5793.

7. Meidan, D., Cohen, R., Haber, S. & Barzel, B. An alternating lock-down strategy for sustainable mitigation of COVID-19. arXiv [q-bio.PE] (2020).

8. Bar-On, Y. M., Flamholz, A., Phillips, R. & Milo, R. SARS-CoV-2 (COVID-19) by the numbers. Elife 9, (2020).

9. Li, R. et al. Substantial undocumented infection facilitates the rapid dissemination of novel coronavirus (SARS-CoV2). Science (2020) doi: 10.1126/science.abb3221.

10. He, X. et al. Temporal dynamics in viral shedding and transmissibility of COVID-19. Infectious Diseases (except HIV/AIDS) (2020) doi: 10.1101/2020.03.15.20036707.

11. Wang, C. et al. Evolving Epidemiology and Impact of Non-pharmaceutical Interventions on the Outbreak of Coronavirus Disease 2019 in Wuhan, China. Epidemiology (2020) doi: 10.1101/2020.03.03.20030593.

12. Leung, K., Wu, J. T., Liu, D. & Leung, G. M. First-wave COVID-19 transmissibility and severity in China outside Hubei after control measures, and second-wave scenario planning: a modelling impact assessment. Lancet (2020) doi: 10.1016/S0140-6736(20)30746-7.

13. Flaxman, S., Mishra, S., Gandy, A. & Others. Estimating the number of infections and the impact of non-pharmaceutical interventions on COVID-19 in 11 European countries. Imperial College preprint (2020).

14. Estimating the number of infections and the impact of nonpharmaceutical interventions on COVID-19 in 14 European countries - Imperial College London. https://imperialcollegelondon.github.io/covid19estimates/#/.

15. Linnarsson, S. To stop COVID-19, test everyone. Medium https://medium.com/@sten.linnarsson/to-stop-covid-19-test-everyone-373fd80eb03b (2020).

16. Roelfs, D. J., Shor, E., Davidson, K. W. & Schwartz, J. E. Losing life and livelihood: a systematic review and meta-analysis of unemployment and all-cause mortality. Soc. Sci. Med. 72, 840–854 (2011).

17. Champredon, D., Dushoff, J. & Earn, D. Equivalence of the Erlang SEIR epidemic model and the renewal equation. (2018) doi: 10.1101/319574.

18. House, T. & Keeling, M. J. Epidemic prediction and control in clustered populations. J. Theor. Biol. 272, 1–7 (2011).

19. Cui, F. et al. Transmission of pandemic influenza A (H1N1) virus in a train in China. J. Epidemiol. 21, 271–277 (2011).

20. Barro, R., Ursúa, J. & Weng, J. The Coronavirus and the Great Influenza Pandemic: Lessons from the ‘Spanish Flu’ for the Coronavirus’s Potential Effects on Mortality and Economic Activity. (2020) doi: 10.3386/w26866.

21. Fan, V., Jamison, D. & Summers, L. The Inclusive Cost of Pandemic Influenza Risk. (2016) doi: 10.3386/w22137.

22. Blanchard, O. & Summers, L. Fiscal Increasing Returns, Hysteresis, Real Wages and Unemployment. (1986) doi: 10.3386/w2034.

23. O’Shaughnessy, T. Unemployment hysteresis and capacity. (2001).

24. Pissarides, C. A. Loss of Skill During Unemployment and the Persistence of Employment Shocks. The Quarterly Journal of Economics vol. 107 1371–1391 (1992).

25. Guerrieri, V., Lorenzoni, G., Straub, L. & Werning, I. Macroeconomic Implications of COVID-19: Can Negative Supply Shocks Cause Demand Shortages? SSRN Electronic Journal doi: 10.2139/ssrn.3570096.

26. Kaplan, G., Moll, B. & Violante, G. Pandemics According to HANK, work in progress. Slides available at https://benjaminmoll.com/HANK_pandemic/.

27. Case, A. & Deaton, A. Deaths of Despair and the Future of Capitalism. (2020) doi: 10.2307/j.ctvpr7rb2.

